# Real-world cardiovascular effects of liraglutide: transportability analysis of the LEADER trial

**DOI:** 10.1101/2025.05.12.25327466

**Authors:** Kevin Josey, Wenhui Liu, Theodore Warsavage, Morten Medici, Kajsa Kvist, Catherine G. Derington, Jane E.B. Reusch, Debashis Ghosh, Sridharan Raghavan

**Affiliations:** US Department of Veterans Affairs Eastern Colorado Health Care System, Aurora, CO; Department of Biostatistics and Informatics, Colorado School of Public Health, Aurora, CO; Novo Nordisk A/S, Copenhagen, Denmark; Division of Cardiology, Department of Medicine, University of Colorado School of Medicine, Aurora, CO; Adult C Child Center for Outcomes Research C Delivery Science, University of Colorado Anschutz Medical Campus, Aurora, CO; Division of Diabetes, Endocrinology, and Metabolism, Department of Medicine, University of Colorado School of Medicine, Aurora, CO; Division of General Internal Medicine, Department of Medicine, University of Colorado School of Medicine, Aurora, CO

**Author notes:** Corresponding authors: Sridharan Raghavan, MD, PhD Rocky Mountain Regional VA Medical Center 1700 North Wheeling Street Aurora, CO 80045, Kevin Josey, PhD Colorado School of Public Health 13001 East 17^th^ Place Aurora, CO 80045.

## Abstract

Appropriate use of recently approved type 2 diabetes treatments depends on the external validity of landmark randomized clinical trials (RCTs) in real-world populations that may differ from trial participants. This study transported effect estimates from the Liraglutide Effect and Action in Diabetes: Evaluation of Cardiovascular Outcome Results (LEADER) trial, a placebo-controlled RCT of liraglutide on cardiovascular outcomes, onto real-world cohorts within the Veterans Affairs (VA) healthcare system. Risk differences (RDs) in survival outcomes, approximated using pseudo-observations of individual survival probabilities, were estimated with augmented inverse probability weighting after balancing baseline characteristics between RCT and target samples using approximate balancing weights. Transported effects of liraglutide compared with placebo on major adverse cardiovascular events (MACE) and all-cause mortality in veterans (“VA-weighted LEADER”) were larger than, though statistically consistent with, the treatment effects observed in LEADER: MACE RD at 3 years of 4.6% (95% CI, 2.2, 7.0) in VA-weighted LEADER vs 1.6% (0.3, 2.9) in LEADER; all-cause mortality RD at 3 years of 2.9% (0.8, 5.1) in VA-weighted LEADER vs 0.9% (−0.1, 1.9) in LEADER. These estimates of the effects of liraglutide in veterans with diabetes provide real-world evidence that can guide diabetes treatment decisions and formulary policies for a high-risk population underrepresented in RCTs.

## Introduction

Randomized clinical trials (RCTs) provide gold-standard evidence for evaluating medication efficacy but often have uncertain external validity, especially when RCT participants are not representative of potential real-world users of a medication.^1^ This uncertain external validity can complicate treatment decision-making at the point of care and may underlie delays in uptake of new, effective therapies.^1,2^ Furthermore, RCTs often motivate regulatory decisions, treatment guidelines, and medication access policies, particularly for new therapies, resulting in regulatory approval, guidelines, and policies that impact diverse real-world populations based on data from non-representative RCT samples.^3–5^ The disconnect between individuals sampled into RCTs and real-world patient populations is particularly acute in type 2 diabetes, a highly prevalent condition affecting a heterogeneous population with a substantial expansion in treatment options over the last decade. For example, RCTs of glucagon-like peptide-1 receptor agonists (GLP-1RAs), potent agents for the treatment of type 2 diabetes and obesity,^6,7^ with demonstrated cardiovascular benefits,^6^ have consistently underenrolled relevant constituencies of potential medication users, leading to gaps in our understanding about the effectiveness of these drugs in many populations.^8–13^ Moreover, many newer diabetes medications with cardiovascular benefits, including GLP-1RAs, are expensive, making their use and accessibility sensitive to treatment coverage policies.

While the most direct method for assessing medication effects in specific target populations is to conduct an RCT within each population, this strategy is impractical, and often infeasible, due to high costs and extensive time requirements. Observational studies

using real-world data, collected after a medication is in routine use, offer an appealing alternative to RCTs but can be limited by confounding and sampling bias. To address these limitations, transportability analyses have emerged as an alternative, middle-ground solution.^14,15^ Methods for transporting effect estimates integrate RCT and real-world data to estimate what the treatment effect would have been if individuals from the real-world target population had been enrolled in the trial.^16^ These methods preserve the benefits of randomization while utilizing data on potential medication users without requiring real-world uptake. Thus, transportability analyses can contribute evidence of efficacy for new medications in representative populations and inform formulary policies for healthcare payors servicing real-world potential users of a new medication.

The objective of this study was to assess the external validity of the Liraglutide Effect and Action in Diabetes: Evaluation of Cardiovascular Outcome Results (LEADER) study— the first RCT to demonstrate cardiovascular benefits of a GLP-1RA (liraglutide)^17^—in a real-world population underrepresented in trials. We transported the results of the LEADER study^17^ to US Department of Veterans Affairs (VA) users with type 2 diabetes, a real-world patient population comprising a small minority in foundational RCTs of GLP-1RAs,^6,17–19^ despite having higher cardiovascular disease risk than non-veteran US adults with diabetes.^20,21^ Methodologically, we extend transportability methods to survival outcomes using pseudo-observations within a doubly robust framework,^14,22,23^ providing a template for transporting RCT results with time-to-event endpoints to real-world target populations.

## Methods

### Conceptual framework for transportability

Several methodological strategies are commonly employed for effect transportation: (1) weighting methods (inverse probability weighting [IPW]), which adjust for differences between the trial and target VA populations by assigning weights based on the likelihood of trial participation given observed covariates, reweighting the RCT sample to resemble the VA cohort; (2) g-computation, which involves modeling the outcome as a function of treatment and covariates to predict outcomes under different treatment scenarios in the target population; and (3) doubly robust (DR) methods (e.g., augmented IPW), which combine weighting and outcome modeling, achieving robustness by requiring only 1 of these 2 models (propensity for trial inclusion or outcome regression) to be correctly specified. We focus on the latter method as it enables the operationalization of machine learning to provide more robust, data-driven estimates without sacrificing parametric rates of convergence (e.g., the central-limit theorem holds for DR estimators despite using models that do not necessarily confer root-n consistency).^24^

### Pseudo-observations for survival probabilities

Pseudo-observations, as introduced by Andersen and Pohar Perme, provide a method for estimating survival probabilities at a given time using a leave-one-out jackknife approach.^22^ For participant *i* = 1,2, . . ., *n*, and given the trial data where the samples have survival data, let *S*\*(*t*) be the Kaplan–Meier estimator of the survival function at time t. The individual pseudo-observations are defined as *S*^.^ᵢ(*t*) = *n* · *S*\*(*t*) − (*n* − 1) · *S*^*("*i*)^(*t*), where *S*\*(*t*) is the Kaplan–Meier estimate of survival at time t using all data, and *S*^*("*i*)^(*t*) is the Kaplan–Meier estimate of survival at time *t* computed after removing individual *i* from the dataset. These pseudo-observations are analyzed as quasi-continuous outcomes with standard regression methods, avoiding direct modeling of censored time-to-event data. We computed pseudo-observations for individual survival probabilities at 6-month intervals from 6 to 54 months.

### Identification of the transported average treatment e=ect

Let *Z_i_* ∈ {0,1} indicate whether a sample belongs to the trial (*Z_i_* = 1) or to the target (*Z_i_* = 0), *A_i_* ∈ {0,1} be the indicator for treatment with liraglutide, and 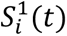 and 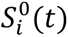 represent the potential survival outcomes under treatment (*A_i_* = 1) and control (*A_i_* = 0), respectively. Here 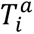 denotes the potential event time under exposure *a* ∈ {0,1}, and 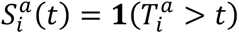 indicates whether unit *i* would remain event-free through follow-up time *t* under exposure *a*, where **1**(⋅) is the indicator function. Under the consistency component of assumption (1) below, the factual event time is 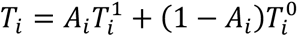, so that the observed survival outcome *S_i_*(*t*) = **1**(*T_i_* > *t*) equals 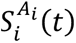. The target parameter defined below is therefore the difference between the corresponding counterfactual survival curves in the target population at a given follow-up time. Define ***X****_i_* = (1, *X_i_*_1_, *X_i_*_2_, …, *X_ip_* B as the set of covariates that we will need to balance and condition our outcome regression upon. The parameter of interest is the target population average treatment effect on survival:

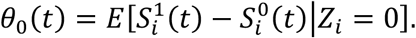

Note that, although the target estimand is defined as a function of follow-up time, in practice we evaluate it at prespecified landmark times (e.g., 36 months) rather than as a continuous curve. To estimate this quantity with the observed data requires 4 assumptions:

(1)Stable unit treatment value assumption, consisting of no interference (the exposure assignment of 1 person does not influence the potential outcome of others) and consistency (the observed outcome corresponds with the factual potential outcome);
(2) Exposure ignorability: 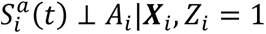, meaning the potential outcomes are independent of exposure when we condition on the confounders, specifically in the trial sample;
(3) Sample exchangeability: the average of the difference between potential outcomes is exchangeable across the trial and the target sample,

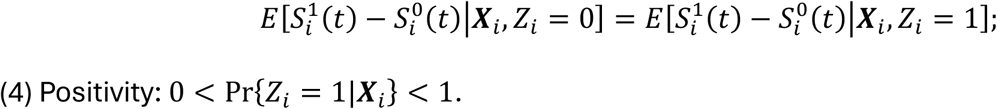
(4) Positivity: 0 < Pr{*Z_i_* = 1|***X****_i_*} < 1.

Positivity requires that every covariate profile in the VA target also appears among trial participants. Sample exchangeability, assumption (3), is the strongest and least testable condition. It fails if a determinant of the treatment effect differs between trial and target and is not captured by the balancing covariates, most plausibly unmeasured background care, adherence, or health behaviors. We assume exchangeability of the conditional treatment effect; the marginal target effect follows by averaging it over the target covariate distribution, and only this difference, not each arm-specific curve, is identified.

### Approximate balancing weights

Approximate balancing weights reweight the trial sample to directly optimize covariate balance while maintaining efficiency. The estimator we implement requires estimated weights *γ_i_*, which are constructed to modify the marginal distribution of covariates ***X****_i_*, particularly the distribution of any effect modifiers, so that the weighted covariate means in the trial sample (*Z_i_* = 1) match those of the target population represented by a target sample (*Z_i_* = 0). We weight the trial sample with approximate balancing weights rather than with inverse-odds weights derived from an estimated model for the probability of trial participation. Modeling and inverting that probability can produce unstable weights when some covariate profiles are rare in the trial, and small modeling errors leave residual imbalance. Balancing weights, on the other hand, solve directly for the covariate balance that transportability requires, improving stability and making the achieved balance transparent.^16,25^

This can be formalized as an optimization problem that identifies the weights that balance the covariate moments^25^ within a standardized range *δⱼ* with

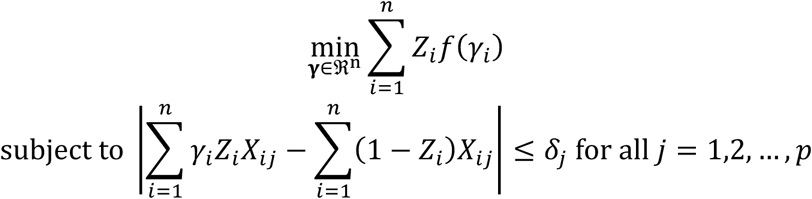

and for a given Bregman distance *f*(*γ_i_*) (e.g., entropy or the squared Euclidean distance).^16,26^ If we select *δ*_j_ = *c* · *σ*_j_where *σ*_j_ is the SD of *X_i_*_j_ in the target population, then we restrict the absolute standardized mean difference (SMD) between the weighted trial covariates and the covariates in the target sample to fall within a constant range *c* of 0. We set *c* = 0.05 to achieve near-exact balance while preserving a larger effective sample size than exact balancing would permit. The effective sample size

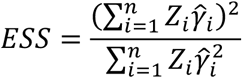

quantifies the information available after weighting. The weights are correctly specified when balancing the covariate means removes the trial-target difference, that is, when participation depends on the covariates through their means. If participation depends on nonlinear functions or interactions, mean balancing need not suffice and the weights are misspecified. Higher-order moments and interactions can be added as further constraints; the nonlinear-misspecification simulation illustrates the bias when such structure is present but only the means are balanced.

### Doubly robust estimation

We constructed DR estimators to estimate treatment effects specific to the target population by transporting average treatment effects from the RCT to the VA target population.^14,23^ The DR augmented inverse-probability-weighted estimator combines both outcome modeling and weighting to correct for potential model misspecifications. The estimated expected treatment effect in the target population (*Z_i_* = 0) at time *t* is given by

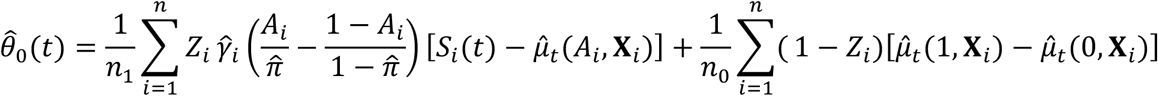

We used Super Learner to fit the outcome model 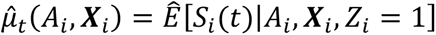 with an ensemble of prediction algorithms incorporating generalized linear model regressions, elastic net regressions, multivariate adaptive regression splines, and random forest regressions.^27^ Nuisance functions were estimated with 5-fold cross-fitting. The outcome model was fitted on the training folds, and the influence function was evaluated on the held-out fold and then averaged. This keeps the remainder second order, so that the influence-function variance remains valid even with flexible, data-adaptive learners. Note that *S_i_*(*t*) is not fully observed under right censoring; in practice, we substitute the jackknife pseudo-observation *S*.*_i_* (*t*) (Appendix S1). By combining the weighting method with an outcome model, this approach remains consistent even if 1 of the 2 components (weights or outcome model) is misspecified.

### Application to LEADER and VA data

Individual-participant LEADER data were shared under a data use agreement with Novo Nordisk A/S; LEADER randomized 9340 patients with type 2 diabetes to liraglutide (1.8 mg daily or maximum tolerated dose) or placebo (median follow-up 3.8 years). Of these, 9336 were included in the analyses. Target-population data were drawn from the VA Corporate Data Warehouse. We mapped LEADER’s major inclusion/exclusion criteria onto the VA data (Table S1) to form a LEADER-eligible cohort (Cohort A: prior atherosclerotic cardiovascular disease [ASCVD], age ≥ 50 years, HbA1c ≥ 7%), additionally excluding individuals with estimated glomerular filtration rate < 15 mL/min/1.73 m², among whom liraglutide prescribing and clinical decision-making differ substantially. Additional cohorts (B–E) relaxed key criteria for sensitivity analyses. Details of data sources, target population definitions, and covariate measurements are provided in **Appendix S1**.

The primary endpoint was time to first major adverse cardiovascular event (MACE), a composite of cardiovascular death, non-fatal myocardial infarction (MI), or non-fatal stroke. Time to non-fatal MI, non-fatal stroke, and all-cause mortality were secondary outcomes. We also evaluated the incidence of selected adverse events. We identified potential effect modifiers and confounders of trial participation: age, self-reported race, measures of diabetes severity (HbA1c, diabetes duration, number of diabetes medications), comorbidities (history of cardiovascular conditions, chronic kidney disease stage, atrial fibrillation, chronic obstructive pulmonary disease [COPD], cancer, liver disease, dementia), and other risk factors (body mass index, hypertension, hyperlipidemia, kidney function, and smoking status). We did not include sex as a covariate in the primary analysis because the VA target population is 97.5% male, creating a near-structural positivity violation. To evaluate differences in the treatment effect between sexes, we conducted a sex-stratified sensitivity analysis.

### Sensitivity analyses

Several sensitivity analyses verified robustness: performing transportability analysis after stratifying both the RCT and target populations by sex; repeating the analysis for alternative target cohorts (B–E), including veterans currently ineligible for GLP-1RA treatment under VA policy; varying modeling parameters—varying balance tolerance, using single outcome learners, and including higher-order and interaction terms when balancing; and a formal unmeasured effect-modifier sensitivity analysis adapted to the transportability setting.^28^ No adjustments for multiplicity were made given the pre-specified primary outcome (composite MACE) and the exploratory nature of secondary outcome and sensitivity analyses.

## Results Simulation study

### Simulation design

Data were generated for a combined sample of *n* = 1000 individuals partitioned into trial (*Z_i_* = 1) and target population (*Z_i_* = 0) subsamples with 2 baseline covariates *X_i_*_1_ ∼ *Normal*(−0.5, 1) and *X_i_*_2_ ∼ *Bernoulli*(0.5). An unmeasured variable *U_i_* ∼*Normal*(0, 1) was included to induce violations of identifying assumptions in specific scenarios. Trial membership was determined by *Z_i_* ∼ *Bernoulli*[*π*(*X_i_*_1_, *X_i_*_2_, *U_i_*)], where *π*(*X_i_*_1_, *X_i_*_2_, *U_i_*) = expit(*α*_0_ + *α*_1_*X_i_*_1_ + *α*_2_*X_i_*_2_ + *α*_3_*U_i_*), with scenario-specific coefficients ***α*** = (*α*_0_, *α*_1_, *α*_2_, *α*_3_). Treatment assignments within the trial followed *A_i_*|*Z_i_* = 1 ∼*Bernoulli*(0.5), reflecting 1-to-1 randomization.

Potential event times were generated from a Weibull proportional hazards model with shape parameter *ν* = 1.5 and baseline hazard *λ*_0_ = 0.2. Specifically, potential event times were obtained by inverting the Weibull survival function

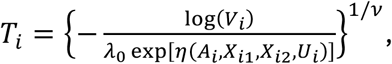

where *V_i_* ∼ *Uniform*(0,1) and the linear predictor incorporated treatment, measured covariates, and scenario-specific terms involving *U_i_*: *η*(*A_i_*, *X_i_*_1_, *X_i_*_2_, *U_i_*) = *β*_1_*X_i_*_1_ + *β*_2_*X_i_*_2_ +*β*_3_*A_i_* + *β*_D_*U_i_* + *β*_E_*A_i_* × *U_i_*. Censoring times *C_i_* were generated independently from an exponential distribution with rate exp(−3). The observed event time was defined as *T̃_i_* = min(*T_i_*, *C_i_*) with event indicator *D_i_* = **1**(*T_i_* ≤ *C_i_*). For individuals in the target population (*Z_i_* = 0), treatment assignment and survival outcomes were treated as unobserved, consistent with the transportability framework where only covariate information is available from the target sample.

We considered the survival probability difference between treatment arms as the target estimand at follow-up times *t* = 1, 2, 3, 4, 5, and 6, transported to the target population. Eight scenarios were examined: (1) baseline: both selection and outcome models correctly specified with no unmeasured confounding; (2) incorrect outcome model: the outcome model omits *U_i_*, which influences survival; (3) incorrect selection model: the selection model omits *U_i_*, which influences trial participation; (4) exchangeability violation: both models omit *U_i_*; (5) nonlinear misspecification: both the selection and outcome models include quadratic and covariate-interaction terms that the linear working models omit; (6) practical positivity violation: extreme coefficients (*α*₁ =2, *α*₂ = −2) inducing near-deterministic selection probabilities for some covariate strata; (7) structural positivity violation: individuals with *X_i_*_1_ > −0.5 were deterministically excluded from the trial (*Z_i_* = 0); and (8) outcome extrapolation: the target covariate distribution is shifted toward larger values of the first covariate, beyond the trial’s support, and the true outcome model adds a hinge term, so that valid estimation requires extrapolation into a region the trial does not cover.

For the simulation study, the DR estimator was implemented using stable balancing weights with *f*(*γ_i_*) being the squared Euclidean distance,^25^ and performance was assessed over 1000 replications using bias (average estimate minus true parameter value) and 95% CI coverage probability. The outcome model was a Gaussian generalized linear regression of the survival pseudo-observations on the covariates and the treatment indicator, using the working linear predictor of each scenario, and the balancing constraints matched covariate means within a standardized tolerance of 0.05. Because these parametric working models lie in a Donsker class, the simulation does not use cross-fitting; it documents the operating characteristics of the influence-function-based Wald interval under each pattern of misspecification.

### Simulation results

Figure 1 displays bias and 95% CI coverage across follow-up for all 8 scenarios. The estimator was approximately unbiased with near-nominal coverage under correct specification and single-nuisance misspecification; bias with degraded coverage arose only when both nuisances were misspecified, through the unmeasured exchangeability violation or nonlinear functional form, with coverage falling furthest under the exchangeability violation. The practical-positivity and outcome-extrapolation scenarios remained near nominal, and the structural-positivity scenario showed larger pointwise bias but near-nominal coverage. Per-scenario bias and coverage ranges are reported in **Table S2**.

**Figure 1.**
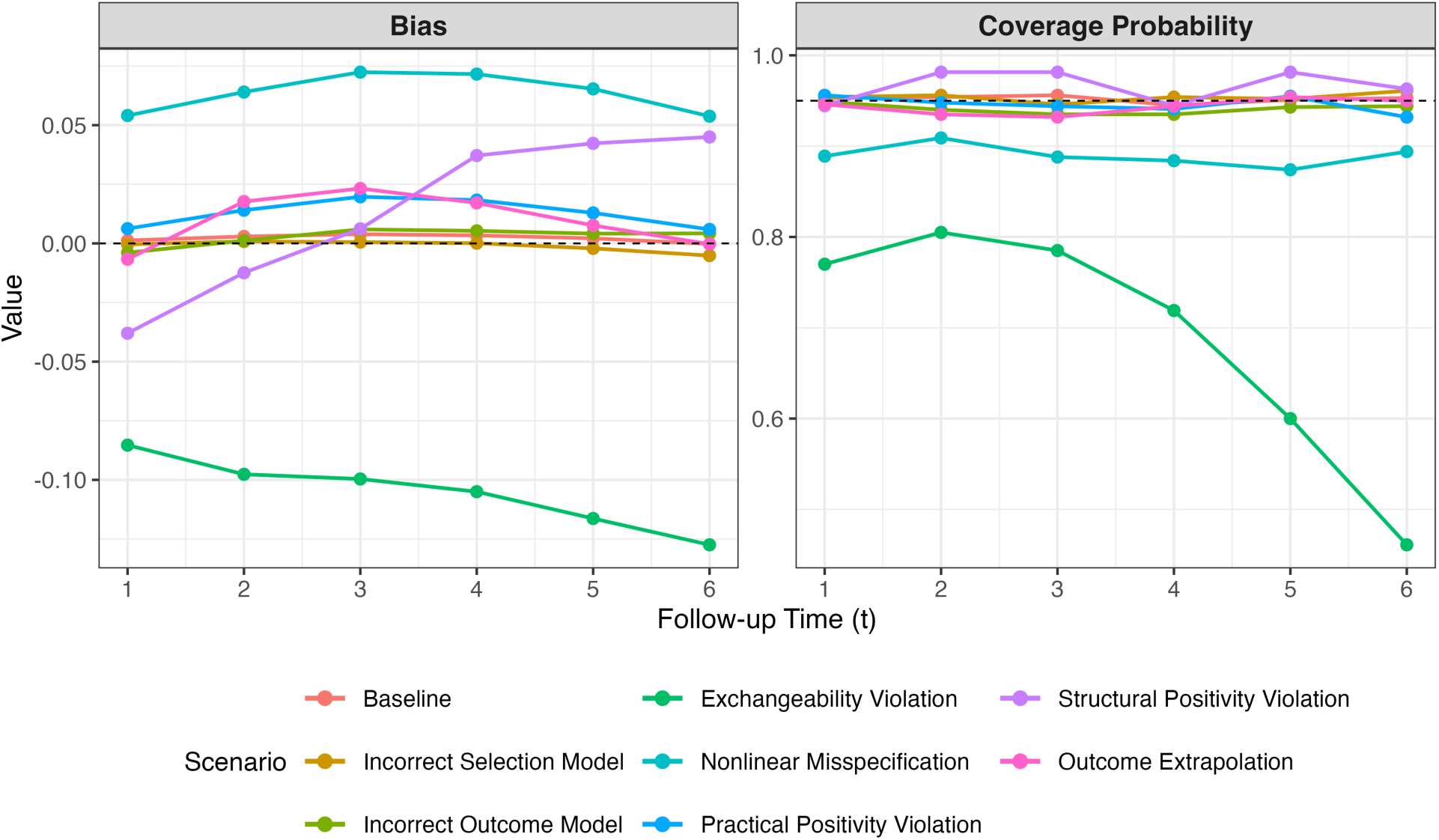
Finite-sample performance of the doubly robust (DR) transportability estimator across simulation scenarios. Bias (**left panel**) and 95% CI coverage probability (**right panel**) are displayed across follow-up times (t = 1, 2, 3, 4, 5, 6) for 8 scenarios: baseline (correct model specification), incorrect outcome model, incorrect selection model, exchangeability violation (both models misspecified), nonlinear misspecification, practical positivity violation, structural positivity violation, and outcome extrapolation. The dashed horizontal lines indicate 0 bias (left) and nominal 95% coverage (right). Results are based on 1000 simulation replications with n = 1000 individuals per replication.

## Results from transporting the LEADER trial

### Baseline characteristics

Cohort A (LEADER-eligible VA cohort) included 357 075 veterans meeting LEADER’s inclusion and exclusion criteria, compared with 9336 participants in the LEADER trial. **Table 1** summarizes baseline characteristics of these 2 groups. Veterans were older on average (mean age of 70.0 years vs 64.3 years in LEADER), predominantly male (97.5% vs 64.3% in LEADER), and were more likely to be Black or African American (14.1% vs 8.3%). Veterans had markedly higher rates of heart failure (21.1% vs 14.0%), atrial fibrillation (16.3% vs 1.9%), COPD (22.3% vs 1.4%), and cancer (36.8% vs 5.9%) compared with LEADER participants. Weight and overlap diagnostics demonstrated partial overlap between trial and target populations (**Table S3**; **Figures S1 and S2**), and, after applying balancing weights, SMDs between the LEADER and VA samples were reduced to ≤0.05 for all balancing variables (**Figure S3**).

**Table 1.** Study participant characteristics at baseline.

|  | **VA Cohort A** | **LEADER** | **P-value** |
| --- | --- | --- | --- |
| Total N | 357 075 | 9336 |  |
| Male sex, n (%) | 348 101 (97.5) | 6001 (64.3) | <.001 |
| Race, n (%) |  |  | <.001 |
| Black or African American | 50 312 (14.1) | 776 (8.3) |  |
| White | 279 341 (78.2) | 7237 (77.5) |  |
| Other | 27 422 (7.7) | 1323 (14.2) |  |
| Age (years), mean (SD) | 70.0 (8.7) | 64.3 (7.2) | <.001 |
| Hemoglobin A1c (%), mean (SD) | 8.8 (1.7) | 8.7 (1.5) | <.001 |
| No. of diabetes medications, n (%) |  |  | <.001 |
| ≤1 | 173 632 (48.6) | 1313 (14.1) |  |
| 2 | 87 554 (24.5) | 3294 (35.3) |  |
| ≥3 | 95 889 (26.9) | 4729 (50.7) |  |
| BMI (kg/m<sup>2</sup>), mean (SD) | 32.6 (6.4) | 32.5 (6.3) | .466 |
| eGFR (mL/min/1.73 m<sup>2</sup>), mean (SD) | 66.4 (21.4) | 79.1 (22.1) | <.001 |
| CAD, n (%) | 196 030 (54.9) | 887 (9.5) | <.001 |
| HF, n (%) | 75 334 (21.1) | 1304 (14.0) | <.001 |
| Prior stroke, n (%) | 85 302 (23.9) | 1506 (16.1) | <.001 |
| Prior MI, n (%) | 44 406 (12.4) | 2862 (30.7) | <.001 |
| Prior revascularization, n (%) | 81 584 (22.8) | 3481 (37.3) | <.001 |
| CKD stage, n (%) |  |  | <.001 |
| Stage 1 | 57 932 (16.2) | 3700 (39.6) |  |
| Stage 2 | 142 760 (40.0) | 3656 (39.2) |  |
| Stage 3 or 4 | 156 383 (43.8) | 1980 (21.2) |  |
| Any cardiovascular disease, n (%) | 303 221 (84.9) | 7731 (82.8) | <.001 |
| Hypertension, n (%) | 306 579 (85.9) | 8508 (91.1) | <.001 |
| Hyperlipidemia, n (%) | 246 848 (69.1) | 7067 (75.7) | <.001 |
| Atrial fibrillation, n (%) | 58 036 (16.3) | 179 (1.9) | <.001 |
| Dementia, n (%) | 19 146 (5.4) | 16 (0.2) | <.001 |
| COPD, n (%) | 79 735 (22.3) | 135 (1.4) | <.001 |
| Cancer, n (%) | 131 325 (36.8) | 551 (5.9) | <.001 |
| Liver disease, n (%) | 26 929 (7.5) | 594 (6.4) | <.001 |
| Smoking status, n (%) |  |  | <.001 |
| Current | 77 955 (21.8) | 1130 (12.1) |  |
| Former | 199 577 (55.9) | 4338 (46.5) |  |
| Never | 79 543 (22.3) | 3868 (41.4) |  |

Abbreviations: BMI, body mass index; CAD, coronary artery disease; CKD, chronic kidney disease; COPD, chronic obstructive pulmonary disease; eGFR, estimated glomerular filtration rate; HF, heart failure; MI, myocardial infarction; VA, Veterans Affairs.

### Outcomes and adverse events

The transported survival probabilities for the primary MACE outcome and secondary outcomes in the primary VA target population were lower for both the liraglutide and placebo arms than in the original LEADER study (**Figure 2, left**). In the VA target population at 36 months, the risk difference (RD) in outcome-free survival favoring liraglutide was 4.6% (95% CI, 2.2%, 7.0%) for composite MACE, 0.9% (−0.4%, 2.2%) for non-fatal stroke, 2.3% (0.6%, 3.9%) for non-fatal MI, and 2.9% (0.8%, 5.1%) for all-cause mortality (**Figure 2, right**). In contrast, in LEADER at 36 months, the RD in outcome-free survival favoring liraglutide was 1.6% (0.3%, 2.9%) for composite MACE and 0.9% (−0.1%, 1.9%) for all-cause mortality. As a complementary summary, we also report the 36-month restricted mean survival time difference (**Table S4**).

**Figure 2.**
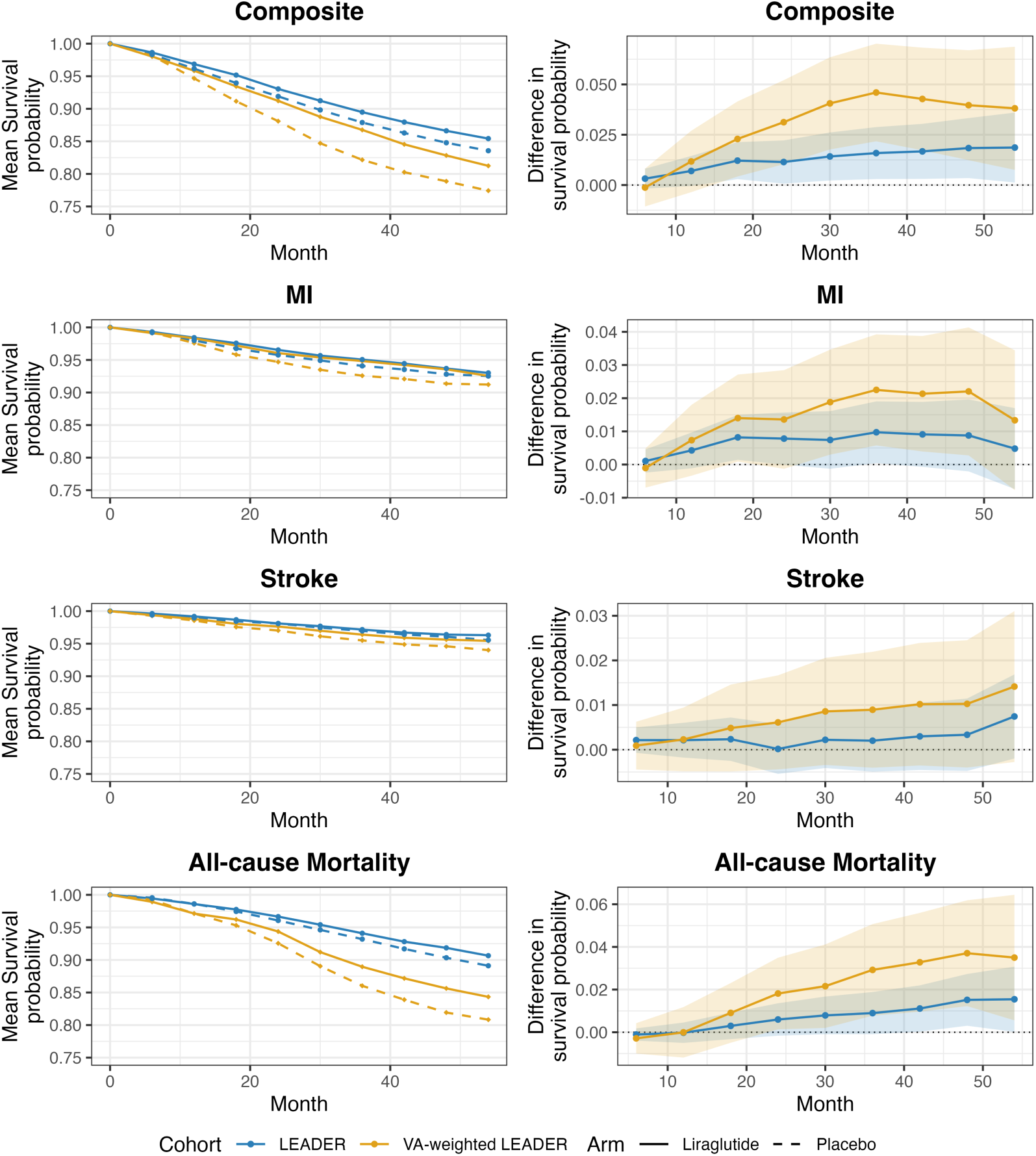
(**Left panel**) Predicted survival probabilities in liraglutide (solid lines) and placebo (dashed lines) arms of the Liraglutide Effect and Action in Diabetes: Evaluation of Cardiovascular Outcome Results (LEADER) trial (blue) and transported to the Veterans Affairs (VA) target population (yellow) for composite major adverse cardiovascular events (composite), non-fatal myocardial infarction (MI), non-fatal stroke, and all-cause mortality. Under assumption (3), only the difference between the arm-specific curves is identified in the target population; the arm-specific survival curves are shown as a decomposition of the identified risk difference rather than as separately identified counterfactual survival probabilities. (**Right panel**) absolute risk differences in event-free survival (liraglutide minus placebo) for composite major adverse cardiovascular events (composite), non-fatal MI, non-fatal stroke, and all-cause mortality in LEADER (blue) and transported to the VA target population (yellow); 95% CIs are shown with colored shading.

To assess external validity in the VA target population of safety outcomes assessed in LEADER, we estimated transported effects of liraglutide vs placebo on selected adverse events. As with the primary and secondary efficacy outcomes, the transported and LEADER 95% CIs overlapped for all adverse events examined (**Figure 3**). The transported RDs were elevated for adverse events leading to discontinuation and for abdominal symptoms, and null for pancreatitis and cancer, each concordant with LEADER. The transported RDs for acute gallbladder disease were imprecise, with 95% CIs that included zero (**Figure 3**, **Table S5**).

**Figure 3.**
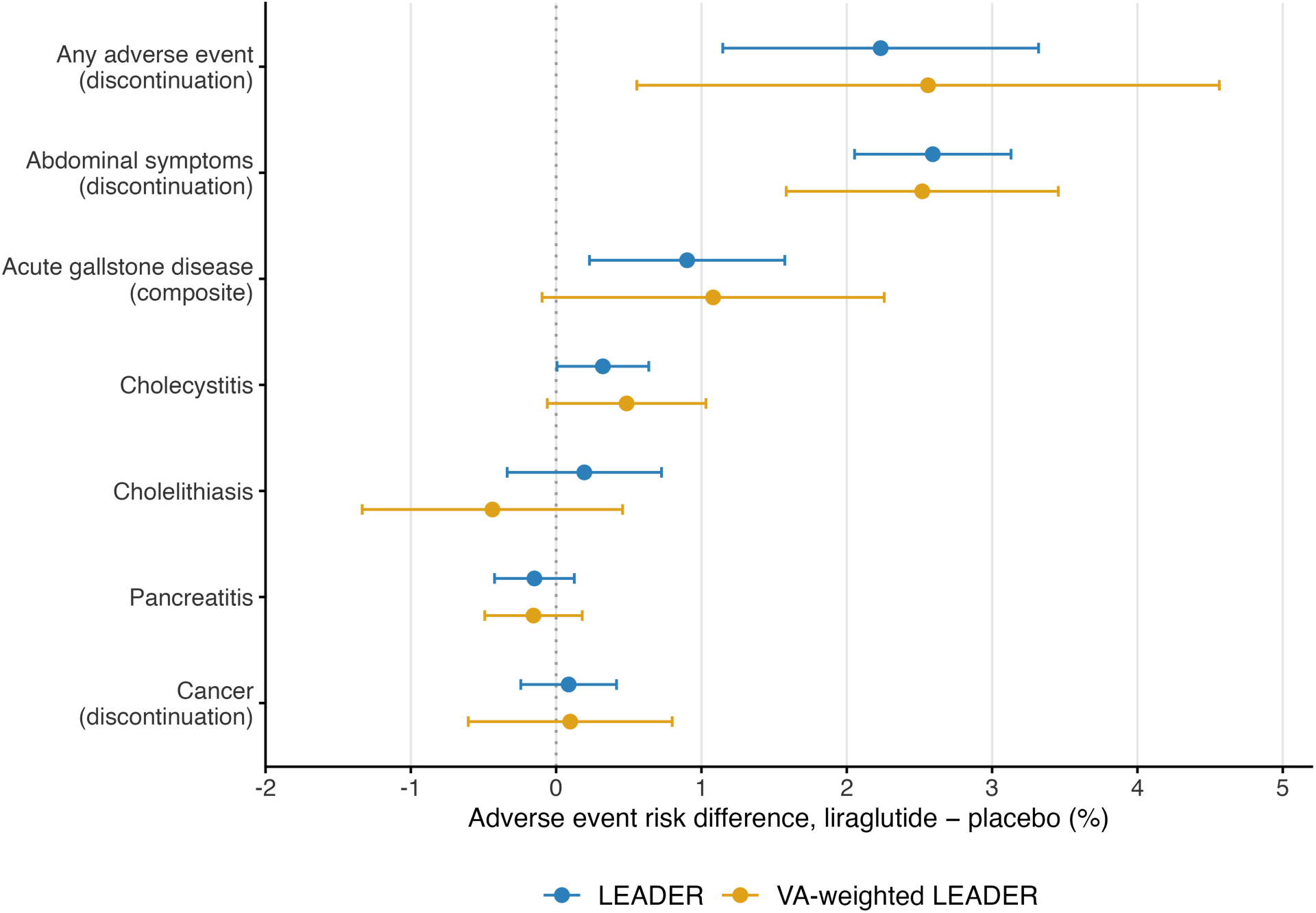
Absolute risk differences at the end of follow-up and 95% CIs for several adverse events recorded in the Liraglutide Effect and Action in Diabetes: Evaluation of Cardiovascular Outcome Results (LEADER) trial (blue) and transported to the Veterans Affairs (VA) target population (yellow). Risk differences greater than 0 (vertical dotted line) reflect greater risk of adverse events in individuals randomized to liraglutide compared to placebo.

### Sensitivity analyses

Baseline characteristics of the additional VA target populations are shown in **Table S6**. In all VA target populations, the CIs of the transported effect estimates overlapped those of LEADER with consistently larger absolute risk reductions for the composite MACE and all-cause mortality outcomes (**Figure 4**). The RDs of the primary and secondary efficacy outcomes were attenuated in the 2 VA target populations that did not require pre-existing ASCVD as an inclusion criterion (Cohorts D and E). Additional robustness analyses appear in the Supplement and were stable throughout: varying the balance tolerance, using single outcome learners, and omitting cross-fitting (**Table S7**); higher-order and interaction balancing (**Table S8**; **Figure S4**); a sex-stratified analysis (**Table S9**; **Figure S5**); and a formal unmeasured effect-modifier sensitivity analysis (**Figure S6**). In that analysis, the robustness value for the 36-month composite MACE effect was 0.07.

**Figure 4.**
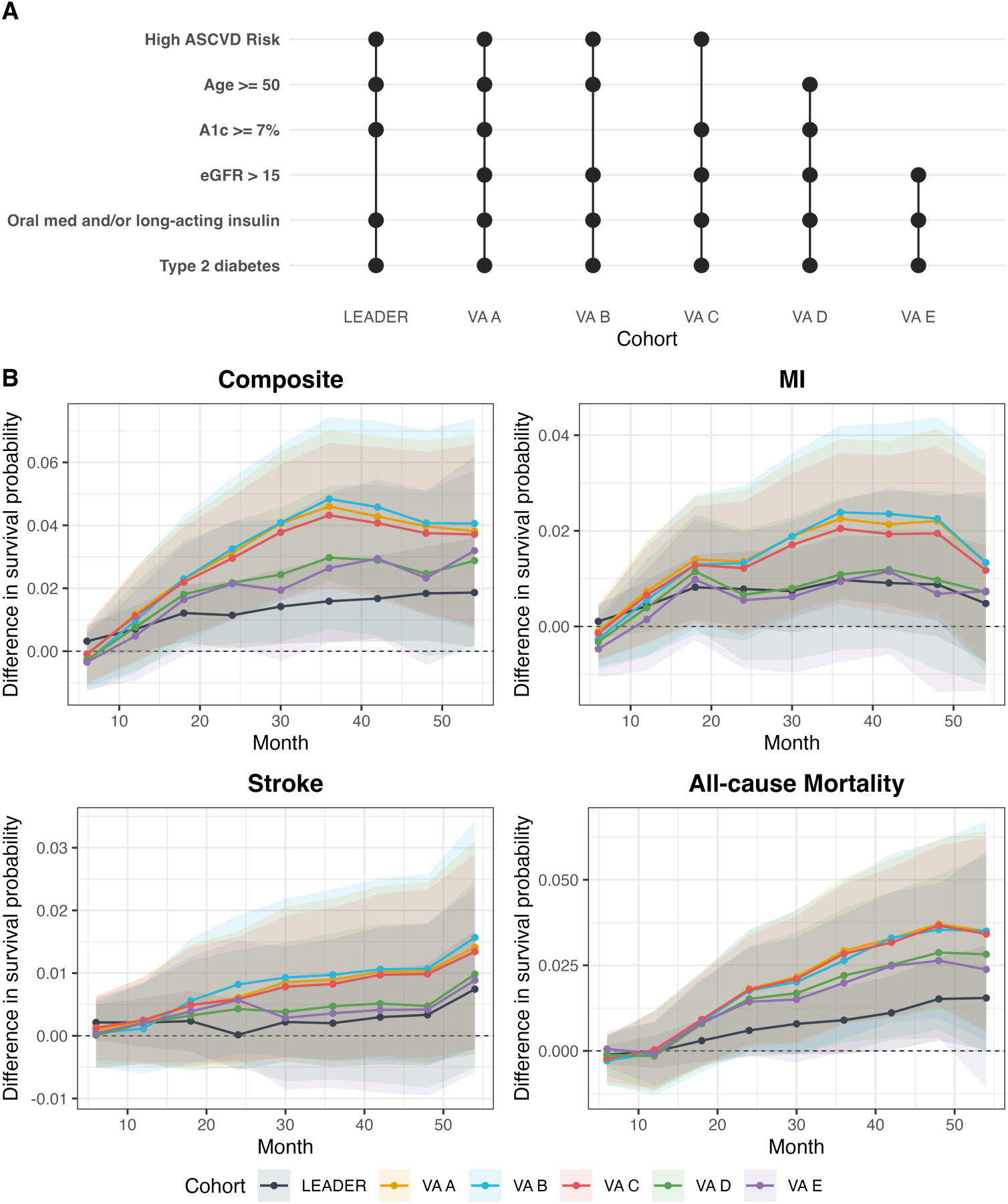
Comparison of treatment effects in the Liraglutide Effect and Action in Diabetes: Evaluation of Cardiovascular Outcome Results (LEADER) trial and LEADER transported to a series of Veterans Affairs (VA) target populations with varying inclusion/exclusion criteria. (**A**) Key inclusion criteria for LEADER and for each VA target population included in the analysis. (**B**) Transported treatment effect estimated as survival probability differences for composite major adverse cardiovascular events (composite), non-fatal myocardial infarction (MI), non-fatal stroke, and all-cause mortality; 95% CIs are shown with colored shading for each transported treatment effect estimate. VA A through VA E indicate results after weighting LEADER to the corresponding target population. Abbreviations: ASCVD, atherosclerotic cardiovascular disease; A1c, hemoglobin A1c; eGFR, estimated glomerular filtration rate calculated using the modification of diet in renal disease equation (units of mL/min/1.73 m^2^).

## Discussion

We found that the cardiovascular effects of liraglutide examined in the LEADER trial transported to a real-world cohort of US veterans with type 2 diabetes. In this population, liraglutide was associated with reduced risk of composite MACE and all-cause mortality with larger absolute benefit than observed in the trial. The transported effect estimates for all outcomes had 95% CIs that overlapped those from the LEADER study. The point estimates for RD of the primary MACE outcome and all-cause mortality were larger when transported to veterans, corresponding to lower numbers needed to treat (NNT) than observed in LEADER (NNT of 22 vs 63 for MACE and 34 vs 111 for all-cause mortality over 3 years). Moreover, the sensitivity analysis in broader VA target populations suggests liraglutide may confer benefit outside the trial inclusion/exclusion parameters and outside the current criteria for use of GLP-1RAs in the VA, though the potential excess benefits of liraglutide in veterans were attenuated when the target population was extended to include individuals without pre-existing ASCVD. That the transported point estimates exceeded the trial’s confidence limits, even as the intervals overlapped, is itself informative. The large baseline differences between LEADER and veterans (Table 1) appear to modify the absolute treatment effect. Because veterans carry higher baseline event risk, a comparable relative effect mathematically produces a larger absolute benefit, and consequently, a lower number needed to treat. We therefore read the transported estimates as evidence that the absolute benefit of liraglutide is larger in this higher-risk population, while cautioning that the limited effective sample size leaves the difference imprecise.

Our findings extend prior real-world replications of GLP-1RA cardiovascular outcome trials. The benefit in Cohorts D and E irrespective of prior ASCVD mirrors real-world studies of GLP-1RAs for ASCVD primary prevention and RCT subgroup analyses showing no heterogeneity by ASCVD history.^6,29^ Our results also support a recent systematic review that found concordance between RCTs and real-world studies for GLP- 1RAs,^30^ despite real-world patients with diabetes infrequently meeting eligibility criteria for GLP-1RA RCTs.^11^ Importantly, transportability analyses explicitly combine RCT data with real-world data unlike prior secondary analyses of RCTs or studies exclusively using real-world data, leveraging the internal validity of the RCT while extending inferences to the target population.

Transportability analyses can inform treatment policy when a healthcare system’s constituents are underrepresented in an RCT. In the VA, formulary decisions often rest on demonstrated outcomes in veterans; our study provides such evidence in the absence of VA-based RCTs.^31,32^ The primary analyses support the use of GLP-1RAs for patients with type 2 diabetes and established ASCVD or high risk of ASCVD. The broader-population sensitivity analyses suggest several LEADER criteria may not be essential for benefit in veterans; because criteria such as prior ASCVD and elevated HbA1c still govern VA GLP- 1RA use, these exploratory results motivate further study of expanded access, balanced against cost, clinical characteristics, patient preferences, and availability. Analogous analyses can evaluate the external validity of other new cardiometabolic therapies; because they require only target-population covariate data, they can generate timely real-world evidence before a medication is taken up in routine care.

Our study has important limitations. First, transportability assumes that any censoring is non-informative or that the censoring mechanisms would be similar in the target population and in the RCT. Moreover, the influence-function-based CIs are guaranteed nominal coverage only when both nuisance models are estimated consistently at sufficiently fast rates. The near-nominal coverage observed in our simulations under single-nuisance misspecification is an empirical finding rather than a general property; coverage degraded when both models were misspecified, as the joint-misspecification scenarios illustrate. Second, while we included many covariates for adjustment, unmeasured differences or effect modifiers between the trial and VA populations could exist. For example, psychosocial factors, diet, exercise, adherence behaviors, or quality of background care might differ and were not captured. Formal unmeasured effect-modifier sensitivity analysis adapted to the transportability setting^28^ (**Figure S6**) indicated that an unmeasured effect modifier would need to be stronger than any measured covariate to attenuate the transported MACE benefit to the null. Third, the VA population is predominantly older men with extensive comorbidities, so our conclusions apply most strongly to VA and similar populations and warrant caution when extrapolating elsewhere. Fourth, the sensitivity analyses examining broader VA target populations (Cohorts B through E) should be interpreted with caution. By relaxing the inclusion criteria that defined LEADER eligibility—such as age, HbA1c, or ASCVD history—these analyses inherently extrapolate to covariate regions where the trial provides no direct information, representing a structural positivity violation. The consistent direction across these populations is suggestive but cannot establish efficacy outside trial-eligible populations; it is hypothesis-generating evidence that the transported effects are not driven solely by the eligibility criteria. Finally, power is governed by the trial size and the variation in weights: the effective sample size relevant to the primary VA target was only about 3472 of the 9336 LEADER participants, more than one-third, and was generally lower for broader populations (**Figure S7**).

By transporting the LEADER trial results to a real-world population, we found that liraglutide’s cardiovascular and mortality benefits appear to extend, on the basis of the measured effect modifiers, to US adults with type 2 diabetes receiving care in the VA, the largest integrated healthcare system in the United States. The results support the use of liraglutide to mitigate cardiovascular disease risk in a real-world population that was poorly represented in the RCT. As healthcare systems increasingly rely on real-world evidence to inform practice and policy, the approach applied here—leveraging robust RCT results and tailoring them to a target population—can serve as a model for evaluating external validity of RCTs of other new therapies to inform medication access policies.

## Data Availability

Code implementing the transportability pipeline (approximate balancing weights, pseudo-observation computation, the Super Learner outcome model, and the doubly robust estimator) is publicly available, as is the code to replicate the simulation study, at https://github.com/kjosey/leader-transport. A deidentified, anonymized limited data set derived from the datasets used for the analysis can be made available upon reasonable request from researchers with necessary human subjects research oversight and in accordance with VA data sharing policies.

## Acknowledgements

Author contributions

K.J., S.R., W.L., and D.G. were responsible for conceptualization, and K.J. and D.G. for methodology. M.M. and K.K. provided the LEADER data as resources and contributed to data curation and validation. W.L. and T.W. contributed to data curation of the VA analytic dataset, and K.J., W.L., and T.W. carried out the formal analysis. K.J. was responsible for software and visualization. S.R. was responsible for funding acquisition, project administration, and supervision. K.J. and S.R. wrote the original draft, and all authors contributed to the writing, review, and editing of the manuscript.

## Data availability

Code implementing the transportability pipeline (approximate balancing weights, pseudo-observation computation, the Super Learner outcome model, and the doubly robust estimator) is publicly available, as is the code to replicate the simulation study, at https://github.com/kjosey/leader-transport. A deidentified, anonymized limited data set derived from the datasets used for the analysis can be made available upon reasonable request from researchers with necessary human subjects research oversight and in accordance with VA data sharing policies.

## Confficts of interest

M.M. and K.K. are employees of Novo Nordisk and provided data access and assistance with reproducing analyses of the original LEADER study. They provided critical feedback on the manuscript but did not shape or modify the reported results, interpretation, or conclusions. The remaining authors report no conflicts of interest.

