## supplement for "Real-world cardiovascular effects of liraglutide: transportability analysis of the LEADER trial"

**ONLINE SUPPLEMENTAL MATERIAL**

**Table of Contents**

Appendix S1. Supplemental Methods
Data Sources
Target Population Definitions
Covariates
Pseudo-observation surrogate
Cross-fitting and variance estimation
Appendix S2. Supplemental Simulation Study
Supplemental Results
Table S1. Inclusion/exclusion criteria mapping
Table S2. Simulation bias and 95% CI coverage in Supplemental Simulation Study
Table S3. Approximate balancing weight diagnostics by cohort
Figure S1. Distribution of the balancing weights
Figure S2. Trial-participation overlap
Figure S3. Covariate balance plots
Table S4. Transported restricted mean survival time differences
Table S5. Transported adverse-event risk differences, Cohort A
Table S6. Baseline characteristics across cohorts
Table S7. Outcome-model and balance-tolerance sensitivity
Table S8. Higher-order balancing sensitivity
Figure S4. Higher-order vs first-moment balancing
Table S9. Sex-stratified transported effects
Figure S5. Sex-stratified transported risk differences
Figure S6. Unmeasured effect-modifier sensitivity
Figure S7. Effective sample sizes
Supplemental References

### Appendix S1. Supplemental Methods

*Data Sources*

Individual-participant data from the LEADER study was shared with the study team through a data use agreement with Novo Nordisk A/S. All VA data are derived from the Corporate Data Warehouse (CDW) which records diagnoses, laboratory values, vital signs, and medication use for individuals receiving care at the VA. Veterans with type 2 diabetes were identified using a validated algorithm based on the presence of two or more ICD-9 or ICD-10 diagnosis codes for diabetes within a two-year period starting from January 1, 2002. To ensure the study population demonstrated evidence of active VA care, we required participants to have at least two outpatient encounters including at least one VA primary care provider visit and at least one prescription medication filled through the VA within two years of the first occurrence of a diabetes diagnosis code.

The target population is represented by a cohort of veterans with type 2 diabetes receiving care in the VA between 2015 and 2023, corresponding to years after the initial results of the LEADER trial were first publicly reported. The VA Eastern Colorado Health Care System Research & Development Committee and the Colorado Multiple Institutional Review Board provided human subjects review and approval of the study.

*Target Population Definitions*

We translated the major inclusion and exclusion criteria of the LEADER trial onto the VA data to create a LEADER-eligible VA cohort (**Table S1**). As in LEADER, VA patients could previously or contemporaneously be untreated for diabetes or treated with oral hypoglycemic medications, long-acting insulin, or a combination of these agents. Unlike the LEADER study, however, we excluded individuals with estimated glomerular filtration rate (eGFR) calculated with the Modification of Diet in Renal Disease equation <15 ml/min/1.73m² as liraglutide prescribing patterns and clinical decision-making differ substantially in patients with kidney failure, limiting the relevance of transporting LEADER results to this population.

From these administrative criteria, the primary VA target population ("Cohort A") most closely matches the LEADER study sample, requiring individuals to have a history of atherosclerotic cardiovascular disease (ASCVD), age ≥50 years, and HbA1c ≥7% at baseline. LEADER included individuals with age ≥60 years with at least one cardiovascular risk factor, but this criterion was not mapped to the VA due to uncertainty surrounding the data quality for the cardiovascular risk factors used to define LEADER eligibility.

To examine the potential benefit of liraglutide use in pragmatic populations that were not trial-eligible, additional VA target populations were created for sensitivity analyses, relaxing three inclusion criteria of LEADER:

1. Cohort B: relaxed the HbA1c requirement (included all HbA1c values)
2. Cohort C: removed the age restriction (included age <50 years)

3. Cohort D: did not require a history of ASCVD
4. Cohort E: imposed none of these three criteria

*Covariates*

We identified potential effect modifiers and confounders of trial participation, variables that might differ between the trial and VA populations which also may modify treatment effects: age, self-reported race, measures of diabetes severity (HbA1c, diabetes duration, number of diabetes medications), comorbidities (history of cardiovascular conditions including coronary artery disease, heart failure, prior stroke, prior MI, prior revascularization; chronic kidney disease stage; atrial fibrillation; chronic obstructive pulmonary disease [COPD]; cancer; liver disease; dementia), and other risk factors (body mass index [BMI], blood pressure, lipids, kidney function, smoking status).

We did not include sex as a covariate in the primary analysis because the VA target population is 97.5% male, creating a near-structural positivity violation. Very few female veterans would receive meaningful weight, and balancing on sex would effectively exclude most LEADER female participants from contributing to the transported estimate. Including sex as a balancing covariate would thus transport LEADER primarily to veteran men, which is largely what the unweighted VA sample already represents. We nevertheless conducted a sex-stratified sensitivity analysis, transporting men and women separately, to verify that this decision did not materially affect the transported effect estimates.

The entropy balancing optimization requires that the balancing covariates are linearly independent; we verified this condition was satisfied by confirming that the design matrix of balancing covariates had full column rank prior to weight estimation.

*Pseudo-observation surrogate*

The outcome model regresses the survival pseudo-observations on the covariates and the treatment indicator. The pseudo-observation for individual i at time t is the jackknife quantity obtained from the Kaplan-Meier estimator; under independent censoring its conditional expectation equals the conditional survival probability, so it is asymptotically unbiased for the quantity it replaces (Andersen and Pohar Perme 2010; Graw, Gerds, and Schumacher 2009). Because each pseudo-observation is formed by leaving one observation out, the pseudo-observations are weakly dependent across individuals; this dependence is of smaller order and vanishes with sample size, so the probability limit of the outcome regression is unchanged when the computable pseudo-observation replaces the unobserved survival indicator $S_i(t)$, and the rate of convergence is governed by the regression estimator rather than by the pseudo-observation step (Overgaard, Parner, and Pedersen 2017; Jacobsen and Martinussen 2016). The doubly-robust transported estimator therefore inherits its usual large-sample properties when it is constructed from estimated pseudo-observations.

*Cross-fitting and variance estimation*

For the data analysis the nuisance functions were estimated with five-fold cross-fitting. Within each fold the Super Learner outcome model and the approximate balancing weights were fit on the complementary four folds and the efficient influence function was evaluated on the held-out fold, with the transported estimate and its variance obtained by averaging the fold-specific influence functions. Cross-fitting removes the own-observation overfitting bias that can arise when flexible learners estimate the nuisances and renders the remainder term second order, entering as the product of the errors in the two nuisance models (Chernozhukov et al. 2018). Nominal coverage of the influence-function-based interval is therefore guaranteed when both nuisance models are estimated consistently, with the product of their estimation errors of smaller order than $n^{-1/2}$. If one nuisance model is inconsistent, the point estimator remains consistent by double robustness, but the estimated influence function no longer captures all first-order variation of the estimator, so nominal coverage is not guaranteed. In the simulation, by contrast, the working models are parametric and lie in a Donsker class, so the remainder is controlled without sample splitting and cross-fitting is not required. The near-nominal coverage observed there under single-nuisance misspecification is an empirical finding rather than a guarantee of doubly robust inference; coverage degraded when both nuisance models were misspecified, as the joint-misspecification scenarios illustrate.

Standard errors for the risk difference estimates were computed using influence function-based variance estimators that account for uncertainty in both the weighting and outcome models. No adjustments for multiplicity were made given the pre-specified primary outcome (composite MACE) and the exploratory nature of secondary outcome and sensitivity analyses; confidence intervals for secondary and sensitivity analyses should be interpreted accordingly. All analyses were completed in R v4.4.1 (R Core Team 2025, Vienna, Austria).

**Appendix S2. Supplemental Simulation Study**

The doubly-robust transportability estimator was evaluated over 1,000 replications (n = 1,000 per replication) across eight scenarios spanning correct specification, single- and joint-nuisance misspecification (an omitted variable and nonlinear functional form), practical and structural positivity violations, and outcome extrapolation; the data-generating process and estimand are described in the main-text Simulation Study, and bias and coverage across follow-up are shown in Figure 1. Per-scenario summaries are given in Table S2.

**Supplemental Results**

**Table S1.** Inclusion/exclusion criteria for LEADER study and mapping to VA data.

| LEADER | VA |
|---|---|
| **Baseline date** | **Baseline date** |
| Date of randomization | Earliest date after meeting all below inclusion/exclusion criteria that any new diabetes medication prescription was filled in VA care |
| **Major inclusion criteria** | **Major inclusion criteria** |
| Patients with type 2 diabetes | Patients with type 2 diabetes based on ICD 9 or 10 codes[a] |
| Treatment history:<br>i. No diabetes medications<br>ii. One or more oral antihyperglycemic agents<br>iii. Selected insulins (human neutral protamine Hagedorn, long-acting analogue, premixed)<br>iv. Combination of above | Treatment history from VA pharmacy records and linked data from CMS data:<br>i. No diabetes medications<br>ii. One or more oral antihyperglycemic agents<br>iii. Selected insulins (human neutral protamine Hagedorn, long-acting analogue, premixed)<br>iv. Combination of above |
| HbA1c ≥ 7.0% | HbA1c ≥ 7.0% |
| Age ≥ 50 years with at least one cardiovascular coexisting condition: | Age ≥ 50 years (from VA enrollment records) with at least one cardiovascular coexisting condition[b,c]: |
| i. Coronary heart disease | i. ICD9 or 10 codes or procedure codes for myocardial infarction, coronary artery disease, percutaneous coronary intervention, coronary artery bypass graft |
| ii. Cerebrovascular disease | ii. ICD9 or 10 codes for stroke, transient ischemic attack, cerebrovascular disease |
| iii. Peripheral vascular disease | iii. ICD9 or 10 codes or procedure codes for peripheral vascular disease |
| iv. Chronic kidney disease of stage 3 or greater | iv. Baseline CKD stage 3 or greater based on eGFR <60 ml/min/1.73m$^2$ calculated using MDRD equation<br>OR<br>ICD9 or 10 codes for chronic renal failure |
| v. Chronic heart failure of New York Heart Association class II or III | v. ICD9 or 10 codes for congestive heart failure, systolic heart failure |

| | |
|---|---|
| Age ≥ 60 years with at least one cardiovascular risk factor, as determined by the investigator: Microalbuminuria or proteinuria; hypertension and left ventricular hypertrophy, left ventricular systolic or diastolic dysfunction, or an ankle-brachial index of less than 0.9. | Not mapped due to poor availability of standardized high-quality data related to albuminuria, echocardiographic parameters, ankle brachial index |
| **Major exclusion criteria** | **Major exclusion criteria** |
| Type 1 diabetes | Type 1 diabetes based on ICD9 or 10 codes |
| Use of GLP-1 receptor agonists, dipeptidyl peptidase 4 (DPP-4) inhibitors, pramlintide, or rapid-acting insulin | Treatment history from VA pharmacy records to exclude individuals based on VA prescription for GLP-1RA, DPP-4 inhibitors, pramlintide, or rapid-acting insulin |
| Familial or personal history of multiple endocrine neoplasia type 2 or medullary thyroid cancer | Not mapped |
| Occurrence of an acute coronary or cerebrovascular event within 14 days before screening and randomization | No inpatient or outpatient ICD9 or 10 diagnosis codes for myocardial infarction, stroke, transient ischemic attack |
| | eGFR <15 ml/min/1.73m$^2$ or on dialysis<br>• Additional exclusion criterion due to low numbers in among otherwise eligible individuals and uncertain VA eligibility for GLP-1RA treatment |

[a] Safford et al. *Diabetes Care*. 2004; 27 Suppl 2: B10-21.

[b] Raghavan et al. *Journal of the American Heart Association*. 2019; 8(4): e011295.

[c] Honerlaw et al. *Journal of the American Medical Informatics Association*. 2024; 31(5): 1126-1134.

**Table S2. Finite-sample bias and 95% confidence interval coverage of the doubly-robust transportability estimator, by simulation scenario (ranges across follow-up times t = 1–6).**

| **Scenario** | **Bias (range)** | **95% CI coverage (range)** |
|---|---|---|
| Baseline (correct specification) | 0.000 to 0.004 | 0.95 to 0.96 |
| Incorrect outcome model | −0.004 to 0.006 | 0.94 to 0.95 |
| Incorrect selection model | −0.005 to 0.001 | 0.95 to 0.96 |
| Exchangeability violation (both) | −0.127 to −0.085 | 0.46 to 0.81 |
| Nonlinear misspecification (both) | 0.054 to 0.072 | 0.87 to 0.91 |
| Practical positivity violation | 0.006 to 0.020 | 0.93 to 0.96 |
| Structural positivity violation | −0.038 to 0.045 | 0.94 to 0.98 |
| Outcome extrapolation | −0.007 to 0.023 | 0.93 to 0.95 |

Bias is the mean estimate minus the true transported survival-probability difference. The estimator is approximately unbiased with near-nominal coverage under correct specification and single-nuisance misspecification; bias with degraded coverage arises only when both nuisances are misspecified (the exchangeability and nonlinear scenarios); the structural positivity violation produces bias but retains near-nominal coverage.

**Table S3. Approximate balancing weight diagnostics by VA target cohort.**

| VA cohort | Effective sample size | Mean | Median | SD | Max | 99th pct |
|---|---|---|---|---|---|---|
| A (primary) | 3,472 | 1.00 | 0.44 | 1.30 | 8.10 | 5.25 |
| B | 2,869 | 1.00 | 0.11 | 1.50 | 9.32 | 6.21 |
| C | 3,750 | 1.00 | 0.57 | 1.22 | 7.14 | 4.91 |
| D | 3,159 | 1.00 | 0.28 | 1.40 | 7.95 | 5.64 |
| E | 2,422 | 1.00 | <0.01 | 1.69 | 9.35 | 7.38 |

Weights are normalized to mean 1 within the trial sample (n = 9,336). No weight trimming or truncation was applied. The right-skewed distribution (median far below the mean) indicates that a minority of LEADER participants carry most of the weight, consistent with the modest effective sample sizes.

**Figure S1. Distribution of the approximate balancing weights for the primary VA target population (Cohort A).**

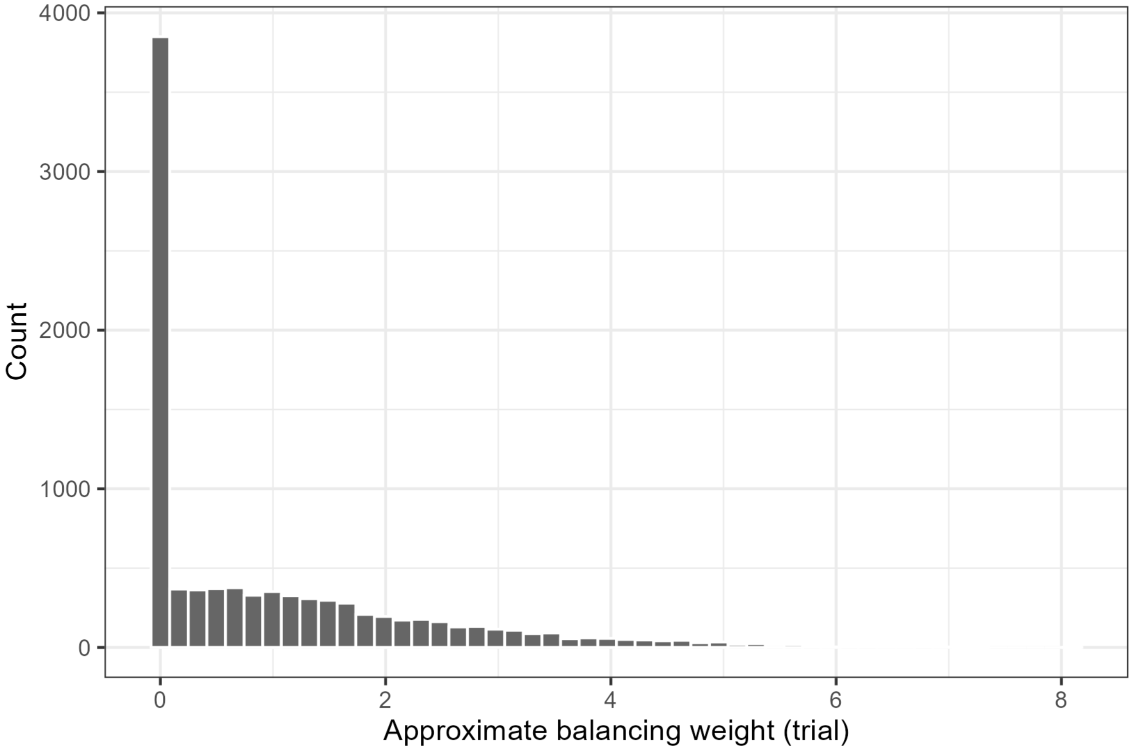


Histogram of the normalized balancing weights assigned to LEADER participants when transporting to Cohort A. No trimming or truncation was applied. The distribution is right-skewed, consistent with an effective sample size of approximately 3,472 of 9,336.

**Figure S2. Overlap in the estimated probability of trial participation between LEADER and the VA target population (Cohort A).**

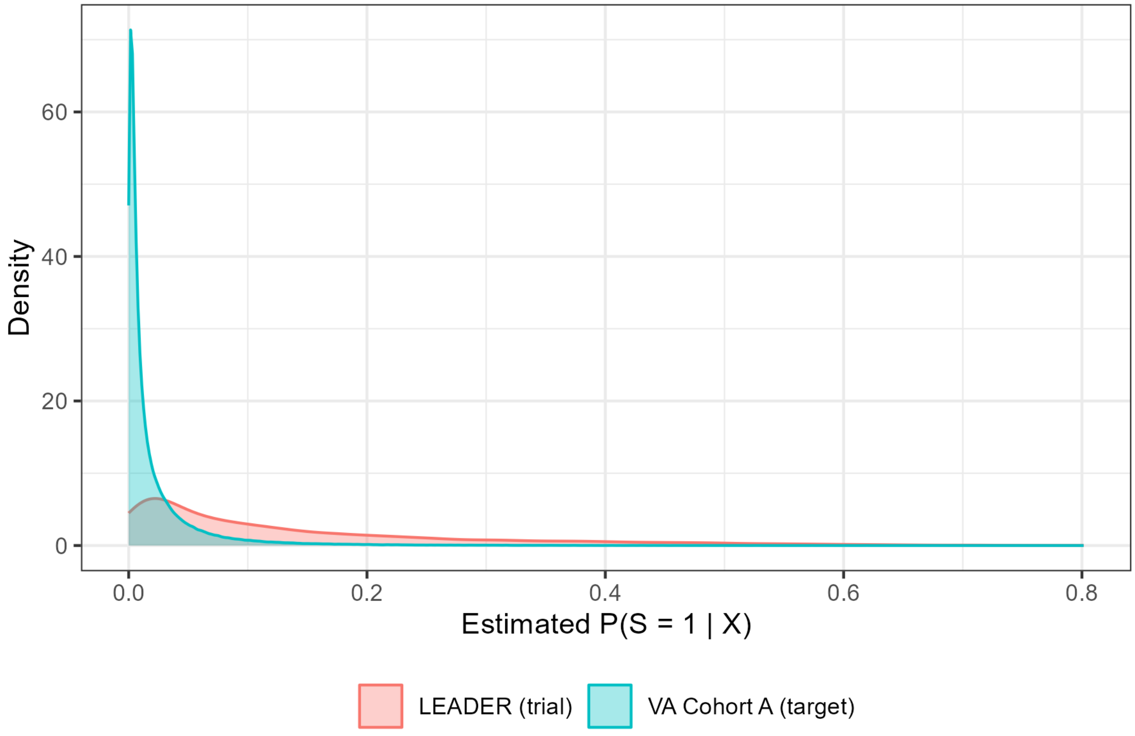


Distributions of the estimated probability of trial participation for LEADER participants and the VA target. Regions of limited overlap indicate where the practical positivity assumption is strained.

**Figure S3.** Covariate balance plots comparing LEADER to VA target populations before and after application of approximate balancing weights.

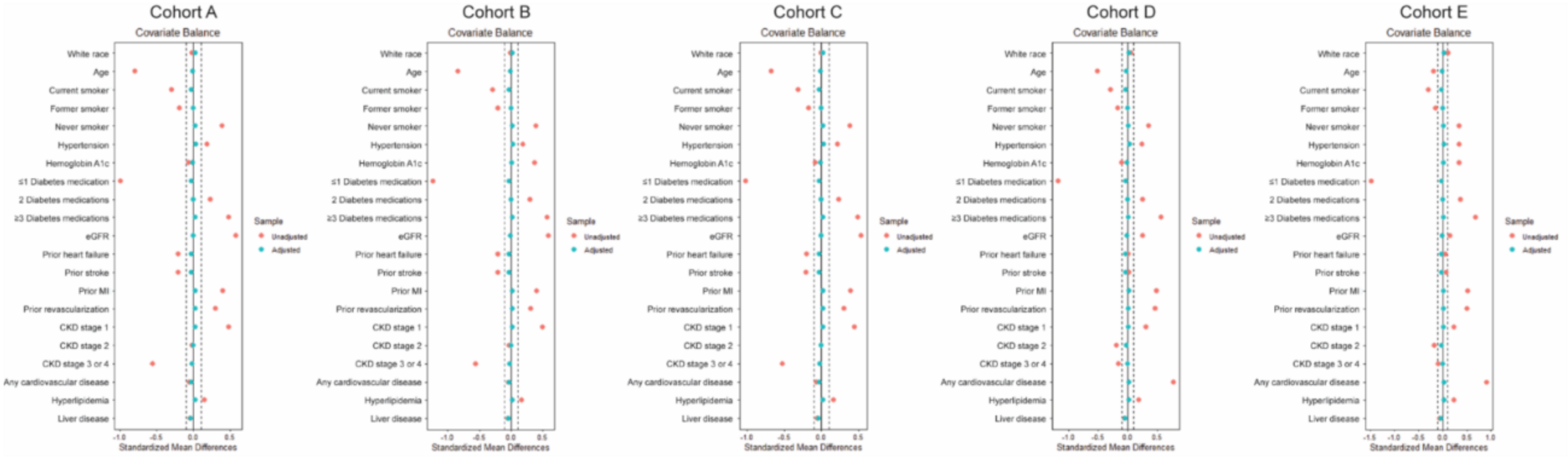


Standardized mean differences (SMD) for baseline covariates before (blue markers) and after (red markers) weighting in each VA cohort (A-E) using approximate balancing weights. The vertical dashed lines (at ±0.1) denote the threshold for negligible imbalance. Baseline differences between the LEADER and VA populations are substantially reduced after weighting, with all SMDs <0.05 after weight application.

**Table S4. Transported 36-month restricted mean survival time (RMST) differences, Cohort A.**

| Outcome | RMST difference at 36 months, months (liraglutide − placebo) |
|---|---|
| Composite MACE | 0.77 |
| Non-fatal MI | 0.38 |
| Non-fatal stroke | 0.16 |
| All-cause mortality | 0.36 |

RMST differences obtained by integrating the transported survival-curve difference from 0 to 36 months. Positive values favor liraglutide.

**Table S5. Transported risk differences for selected adverse events at the end of follow-up, Cohort A.**

| Adverse event | LEADER, RD (95% CI) | VA-weighted LEADER, RD (95% CI) |
|---|---|---|
| Any adverse event leading to discontinuation | 2.23 (1.15, 3.32) | 2.56 (0.56, 4.56) |
| Abdominal symptoms leading to discontinuation | 2.59 (2.05, 3.13) | 2.52 (1.58, 3.46) |
| Acute gallstone disease (composite) | 0.90 (0.23, 1.57) | 1.08 (−0.10, 2.26) |
| Cholecystitis | 0.32 (0.01, 0.64) | 0.49 (−0.06, 1.03) |
| Cholelithiasis | 0.19 (−0.34, 0.73) | −0.44 (−1.33, 0.46) |
| Pancreatitis | −0.15 (−0.42, 0.13) | −0.16 (−0.49, 0.18) |
| Cancer leading to discontinuation | 0.09 (−0.24, 0.42) | 0.10 (−0.60, 0.80) |

RD, risk difference in cumulative incidence (liraglutide − placebo), in percentage points; CI, confidence interval. LEADER estimates are unadjusted trial differences; VA-weighted LEADER estimates are transported to the Cohort A target population using the same approximate balancing weights as the primary analysis. Positive values indicate higher adverse-event risk with liraglutide. These estimates correspond to Figure 3.

**Table S6.** Baseline participant characteristics in LEADER and across VA target populations (Cohorts A-E).

| | LEADER | VA A | p-value | VA B | p-value | VA C | p-value | VA D | p-value | VA E | p-value |
|---|---|---|---|---|---|---|---|---|---|---|---|
| n | 9,336 | 357,075 | | 494,682 | | 370,536 | | 568,311 | | 893,420 | |
| Male sex, n (%) | 6,001 (64.3) | 348,101 (97.5) | <0.001 | 481,991 (97.4) | <0.001 | 360,526 (97.3) | <0.001 | 550,033 (96.8) | <0.001 | 857,093 (95.9) | <0.001 |
| Race, n (%) | | | <0.001 | | <0.001 | | <0.001 | | <0.001 | | <0.001 |
| Black or African American | 776 (8.3) | 50,312 (14.1) | | 71,031 (14.4) | | 53,832 (14.5) | | 95,806 (16.9) | | 163,913 (18.3) | |
| White | 7,237 (77.5) | 279,341 (78.2) | | 385,283 (77.9) | | 287,939 (77.7) | | 425,410 (74.9) | | 651,401 (72.9) | |
| Other | 1,323 (14.2) | 27,422 (7.7) | | 38,368 (7.8) | | 28,765 (7.8) | | 47,095 (8.3) | | 78,106 (8.7) | |
| Age (years), mean (SD) | 64.3 (7.2) | 70.0 (8.7) | <0.001 | 70.3 (8.7) | <0.001 | 69.1 (9.8) | <0.001 | 68.0 (8.9) | <0.001 | 65.7 (11.5) | <0.001 |
| Hemoglobin A1c (%), mean (SD) | 8.7 (1.5) | 8.8 (1.7) | <0.001 | 8.1 (1.8) | <0.001 | 8.8 (1.7) | <0.001 | 8.8 (1.7) | <0.001 | 8.2 (1.9) | <0.001 |
| # of diabetes medications, n (%) | | | <0.001 | | <0.001 | | <0.001 | | <0.001 | | <0.001 |
| ≤1 | 1,313 (14.1) | 173,632 (48.6) | | 281,613 (56.9) | | 183,189 (49.4) | | 312,449 (55.0) | | 585,377 (65.5) | |
| 2 | 3,294 (35.3) | 87,554 (24.5) | | 103,695 (21.0) | | 89,937 (24.3) | | 129,557 (22.8) | | 160,262 (17.9) | |
| ≥3 | 4,729 (50.7) | 95,889 (26.9) | | 109,374 (22.1) | | 97,410 (26.3) | | 126,305 (22.2) | | 147,781 (16.5) | |
| BMI (kg/m$^2$), mean (SD) | 32.5 (6.3) | 32.6 (6.4) | 0.466 | 32.4 (6.4) | 0.215 | 32.7 (6.5) | 0.011 | 32.7 (6.4) | 0.021 | 32.9 (6.5) | <0.001 |
| eGFR (ml/min/1.73m$^2$), mean (SD) | 79.1 (22.1) | 66.4 (21.4) | <0.001 | 66.1 (21.2) | <0.001 | 67.4 (22.0) | <0.001 | 73.4 (21.0) | <0.001 | 75.7 (21.9) | <0.001 |
| CAD, n (%) | 887 (9.5) | 196,030 (54.9) | <0.001 | 271,445 (54.9) | <0.001 | 201,690 (54.4) | <0.001 | 196,030 (34.5) | <0.001 | 279,045 (31.2) | <0.001 |
| HF, n (%) | 1,304 (14.0) | 75,334 (21.1) | <0.001 | 105,025 (21.2) | <0.001 | 77,450 (20.9) | <0.001 | 75,334 (13.3) | 0.046 | 107,882 (12.1) | <0.001 |
| Prior stroke, n (%) | 1,506 (16.1) | 85,302 (23.9) | <0.001 | 116,486 (23.5) | <0.001 | 87,758 (23.7) | <0.001 | 85,302 (15.0) | 0.003 | 119,814 (13.4) | <0.001 |
| Prior MI, n (%) | 2,862 (30.7) | 44,406 (12.4) | <0.001 | 60,408 (12.2) | <0.001 | 46,177 (12.5) | <0.001 | 44,406 (7.8) | <0.001 | 62,788 (7.0) | <0.001 |
| Prior revascularization, n (%) | 3,481 (37.3) | 81,584 (22.8) | <0.001 | 110,898 (22.4) | <0.001 | 83,162 (22.4) | <0.001 | 81,584 (14.4) | <0.001 | 112,934 (12.6) | <0.001 |
| CKD stage, n (%) | | | <0.001 | | <0.001 | | <0.001 | | <0.001 | | <0.001 |

| | | | | | | | | | | |
|---|---|---|---|---|---|---|---|---|---|---|
| Stage 1 | 3,700 (39.6) | 57,932 (16.2) | | 76,367 (15.4) | | 66,076 (17.8) | | 137,732 (24.2) | | 249,322 (27.9) | |
| Stage 2 | 3,656 (39.2) | 142,760 (40.0) | | 199,849 (40.4) | | 146,353 (39.5) | | 274,196 (48.2) | | 423,287 (47.4) | |
| Stage 3 or 4 | 1,980 (21.2) | 156,383 (43.8) | | 218,466 (44.2) | | 158,107 (42.7) | | 156,383 (27.5) | | 220,811 (24.7) | |
| Any cardiovascular disease, n (%) | 7,731 (82.8) | 303,221 (84.9) | <0.001 | 416,681 (84.2) | <0.001 | 315,641 (85.2) | <0.001 | 303,221 (53.4) | <0.001 | 433,270 (48.5) | <0.001 |
| Hypertension, n (%) | 8,508 (91.1) | 306,579 (85.9) | <0.001 | 424,287 (85.8) | <0.001 | 315,309 (85.1) | <0.001 | 478,024 (84.1) | <0.001 | 727,632 (81.4) | <0.001 |
| Hyperlipidemia, n (%) | 7,067 (75.7) | 246,848 (69.1) | <0.001 | 339,547 (68.6) | <0.001 | 254,379 (68.7) | <0.001 | 383,962 (67.6) | <0.001 | 586,671 (65.7) | <0.001 |
| Atrial fibrillation, n (%) | 179 (1.9) | 58,036 (16.3) | <0.001 | 84,367 (17.1) | <0.001 | 58,502 (15.8) | <0.001 | 67,460 (11.9) | <0.001 | 100,482 (11.2) | <0.001 |
| Dementia, n (%) | 16 (0.2) | 19,146 (5.4) | <0.001 | 27,031 (5.5) | <0.001 | 19,224 (5.2) | <0.001 | 22,900 (4.0) | <0.001 | 33,080 (3.7) | <0.001 |
| COPD, n (%) | 135 (1.4) | 79,735 (22.3) | <0.001 | 113,715 (23.0) | <0.001 | 80,846 (21.8) | <0.001 | 102,705 (18.1) | <0.001 | 152,681 (17.1) | <0.001 |
| Cancer, n (%) | 551 (5.9) | 131,325 (36.8) | <0.001 | 182,061 (36.8) | <0.001 | 133,405 (36.0) | <0.001 | 198,533 (34.9) | <0.001 | 293,270 (32.8) | <0.001 |
| Liver disease, n (%) | 594 (6.4) | 26,929 (7.5) | <0.001 | 37,403 (7.6) | <0.001 | 28,415 (7.7) | <0.001 | 43,101 (7.6) | <0.001 | 71,164 (8.0) | <0.001 |
| Smoking status, n (%) | | | <0.001 | | <0.001 | | <0.001 | | <0.001 | | <0.001 |
| Current | 1,130 (12.1) | 77,955 (21.8) | | 106,125 (21.5) | | 82,878 (22.4) | | 122,362 (21.5) | | 195,686 (21.9) | |
| Former | 4,338 (46.5) | 199,577 (55.9) | | 280,102 (56.6) | | 204,111 (55.1) | | 310,955 (54.7) | | 480,631 (53.8) | |
| Never | 3,868 (41.4) | 79,543 (22.3) | | 108,455 (21.9) | | 83,547 (22.5) | | 134,994 (23.8) | | 217,103 (24.3) | |

Abbreviations: BMI, Body mass index; eGFR, estimated glomerular filtration rate; CAD, coronary artery disease; HF, heart failure; MI, myocardial infarction; CKD, chronic kidney disease; COPD, chronic obstructive pulmonary disease

**Table S7. Sensitivity of the transported composite MACE risk difference at 36 months to the outcome-model specification, the balance tolerance, and cross-fitting (Cohort A).**

| **Specification** | **Risk difference at 36 months, % [95% CI]** |
|---|---|
| Outcome model: Super Learner ensemble (primary) | 4.60 [2.17, 7.02] |
| Outcome model: lasso (glmnet) | 4.58 [2.15, 7.00] |
| Outcome model: MARS (earth) | 5.12 [2.69, 7.55] |
| Outcome model: random forest (ranger) | 4.43 [1.97, 6.90] |
| Outcome model: GAM (gam) | 4.56 [2.14, 6.97] |
| Balance tolerance c = 0.005 | 4.70 [1.64, 7.75] |
| Balance tolerance c = 0.010 | 4.74 [1.77, 7.71] |
| Balance tolerance c = 0.050 (primary) | 4.60 [2.17, 7.02] |
| Balance tolerance c = 0.100 | 4.18 [2.11, 6.24] |
| Cross-fitting: five-fold (primary) | 4.60 [2.17, 7.02] |
| Cross-fitting: none (K = 1) | 4.47 [2.13, 6.80] |

The transported composite MACE risk difference was stable across outcome-model specifications and across balance tolerances (range 4.2-5.1%), indicating limited sensitivity of the doubly-robust estimate to these analyst choices. Removing cross-fitting (K = 1) changed the 36-month estimate negligibly (4.47% versus 4.60% with five-fold cross-fitting), indicating that sample splitting does not account for the magnitude of the transported effect. The c = 0.050 row reports the primary cross-fit estimate.

**Table S8. Transported composite MACE risk difference at 36 months under first-moment versus higher-order balancing (Cohort A).**

| Balancing specification | Risk difference at 36 months, % [95% CI] |
|---|---|
| First moments (covariate means; primary) | 4.60 [2.17, 7.02] |
| Higher-order moments and pairwise interactions | 4.77 [2.19, 7.35] |

Adding second-order moments and pairwise covariate interactions to the balancing constraints did not materially change the transported estimate, indicating that first-moment balance is adequate for these covariates.

**Figure S4.** Transported risk differences over follow-up under higher-order and interaction balancing versus first-moment (mean) balancing, Cohort A; LEADER is shown for reference.

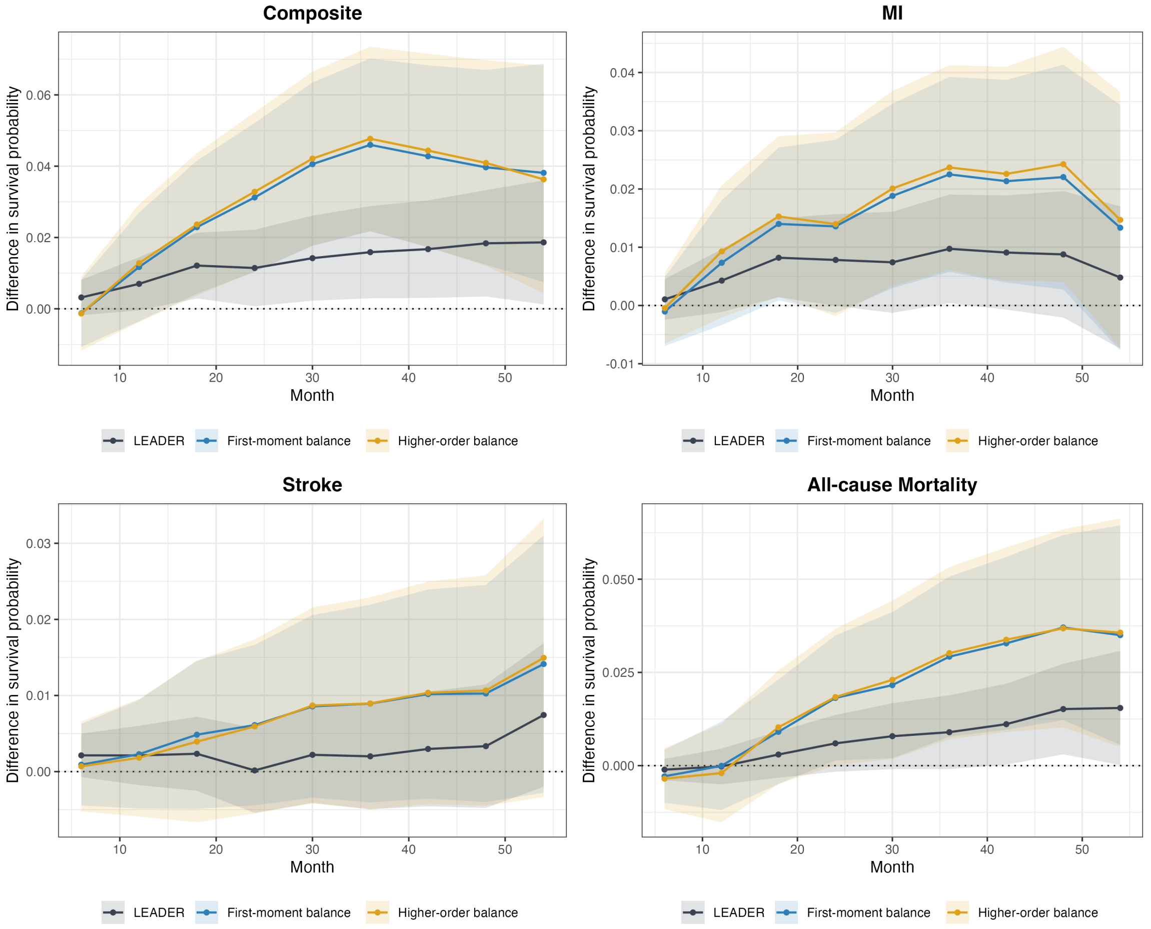


Transported treatment effects estimated as survival probability differences (liraglutide minus placebo) for MACE, non-fatal MI, non-fatal stroke, and all-cause mortality under the primary first-moment (mean) balancing (blue) and under balancing that additionally constrains second-order moments and pairwise interactions (orange), with LEADER shown for reference (grey). Adding higher-order and interaction constraints leaves the transported estimates essentially unchanged.

**Table S9. Sex-stratified transported risk differences at 36 months (Cohort A).**

| Outcome | Men, % [95% CI] | Women (exploratory), % [95% CI] |
|---|---|---|
| Composite MACE | 4.08 [1.07, 7.09] | 3.50 [−0.09, 7.10] |
| All-cause mortality | 2.09 [−0.55, 4.74] | 4.13 [0.81, 7.45] |

Men from LEADER were transported to the male VA population (97.5% of the target); women from LEADER were transported to the small female VA subgroup. The female stratum is small, so the women's estimates are exploratory and imprecise. Transported effects favored liraglutide in both strata.

**Figure S5.** Sex-stratified transported risk differences over follow-up, Cohort A. LEADER men are transported to the male VA target and LEADER women to the small (exploratory) female VA subgroup; LEADER overall is shown for reference.

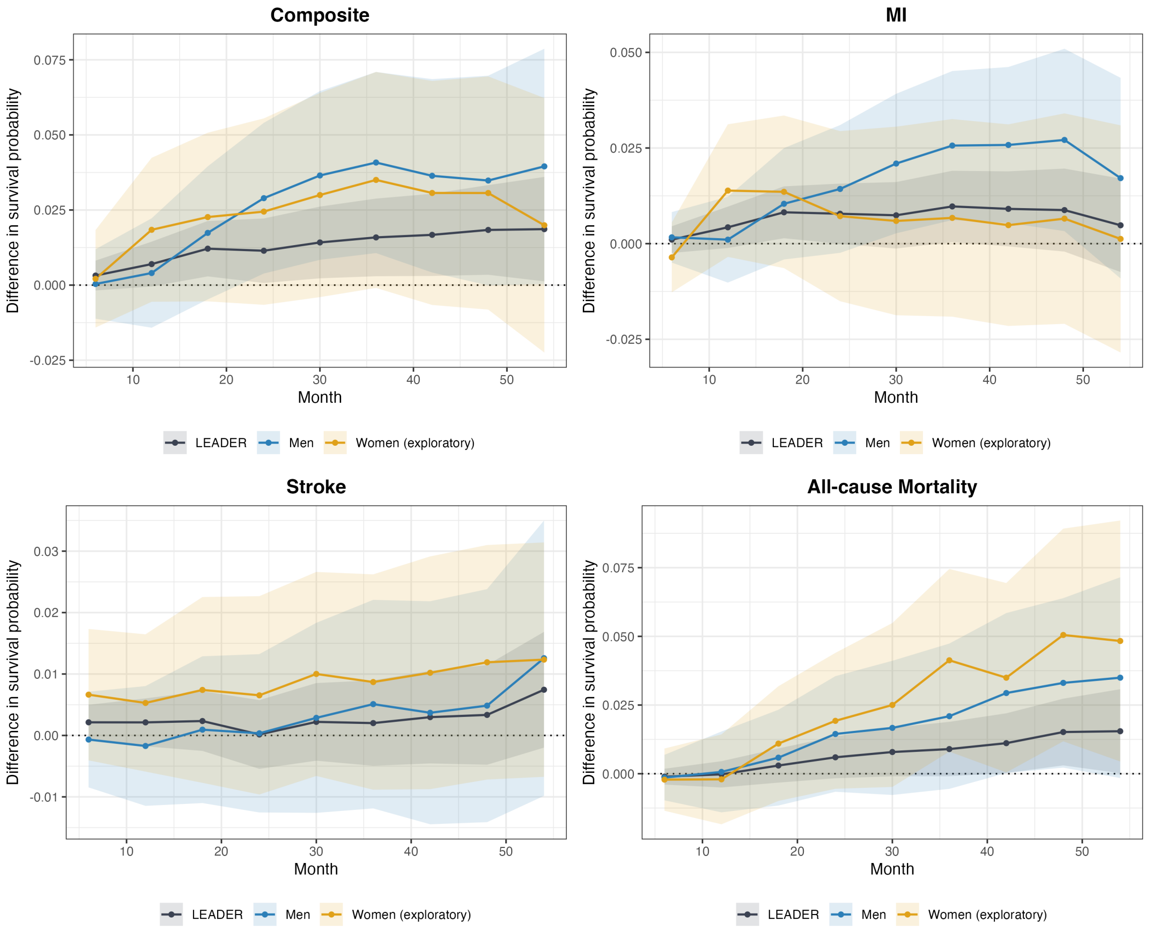


Transported treatment effects estimated as survival probability differences (liraglutide minus placebo) for MACE, non-fatal MI, non-fatal stroke, and all-cause mortality, transported separately by sex: LEADER men to the male VA target population (blue) and LEADER women to the small, exploratory female VA subgroup (orange), with LEADER overall shown for reference (grey). The transported effect favored liraglutide in both strata; the female estimates are imprecise given the predominantly male (97.5%) VA target and are presented as exploratory.

**Figure S6. Sensitivity of the transported 36-month composite MACE effect to an unmeasured effect modifier.**

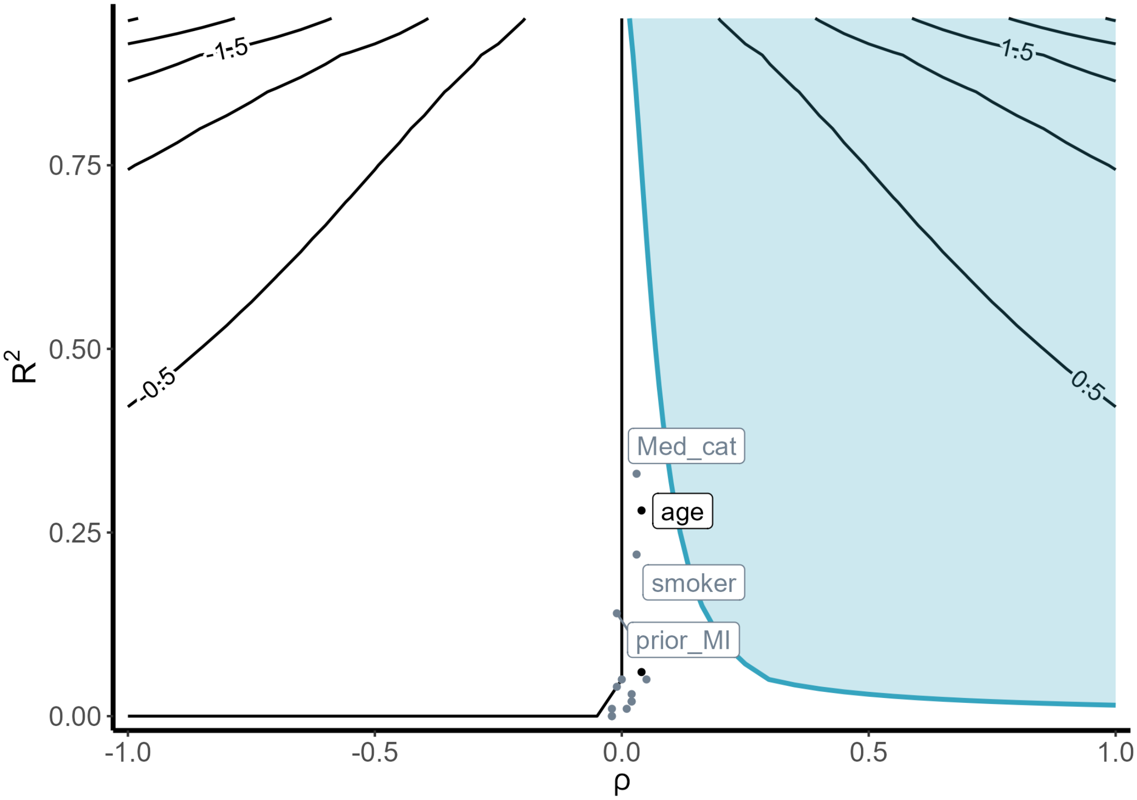


Bias-factor sensitivity analysis adapted to the transportability setting (Huang 2024) for the transported 36-month composite MACE effect. The horizontal axis (ρ) indexes how strongly an unmeasured effect modifier's imbalance between the trial and target populations is related to the outcome, and the vertical axis ($R^2$) indexes the magnitude of that imbalance; together they determine the bias the modifier would induce. Black contour lines give that bias, and the blue contour marks where it becomes large enough to reduce the transported effect exactly to zero; within the blue shaded region beyond this contour, an unmeasured effect modifier would attenuate the transported effect past the null or reverse its sign. Points mark measured covariates positioned by formal benchmarking, i.e., the (ρ, $R^2$) that an omitted modifier as influential as each covariate would induce; the highlighted reference covariates (age and HbA1c) are shown in black and the remaining covariates in grey. The robustness value for the 36-month composite MACE effect was approximately 0.07. An unmeasured effect modifier would need associations with both trial selection and the treatment effect exceeding this value to attenuate the transported effect to the null. All benchmarked covariates fall well outside the shaded region, indicating that no measured covariate reaches the strength required to overturn the result.

**Figure S7.** Effective sample sizes when transporting LEADER to nested VA target populations.

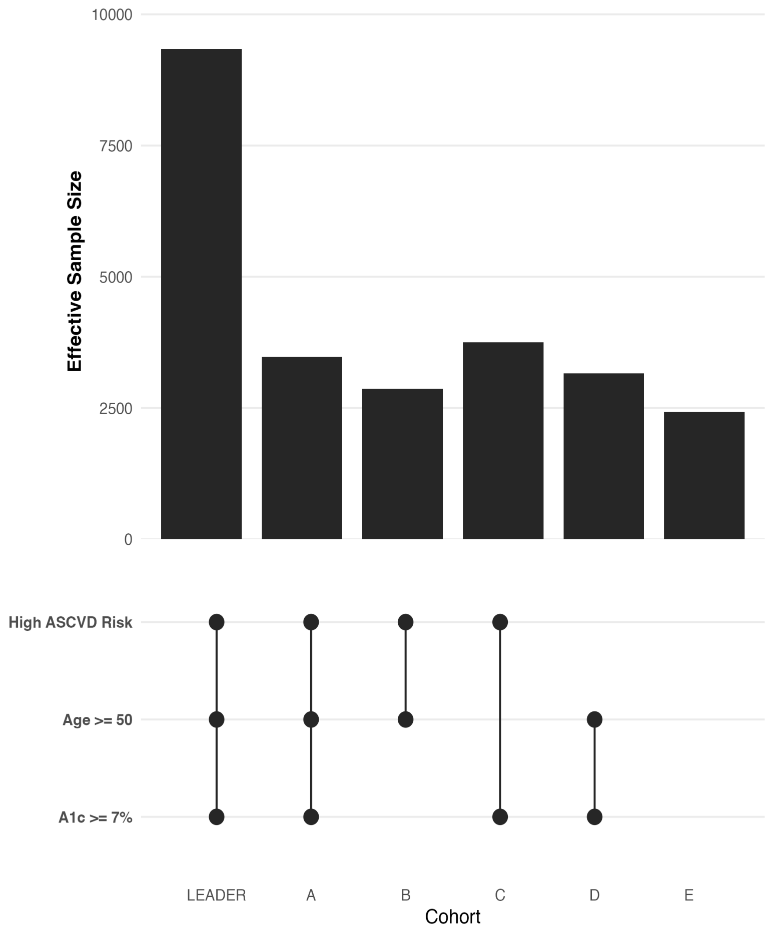


Comparison of inclusion criteria from LEADER and definition of different VA target populations (Cohorts A-E), with effective sample sizes when weighting LEADER data to transport to each cohort. The effective sample size for the primary target population (Cohort A) was approximately 3,472 of 9,336 LEADER participants, indicating that more than one-third of trial participants were similar enough to veterans to contribute meaningful information to the transported effect estimate. Effective sample sizes remain modest across the broader target populations (Cohorts B-E) that relax LEADER eligibility criteria.
